# The effect of job strain and worksite social support on reported side effects of COVID-19 vaccine: a prospective study of employees in Japan

**DOI:** 10.1101/2022.02.24.22271484

**Authors:** Natsu Sasaki, Reiko Kuroda, Kanami Tsuno, Kotaro Imamura, Norito Kawakami

## Abstract

**Objectives:** This prospective study aimed to examine the association of job demands, job control, and the lack of supervisor and coworker support with side effects after receiving COVID-19 vaccination in a sample of employees in Japan.

**Methods:** The data were retrieved from an online panel of full-time employees (E- COCO- J). The analysis included participants who were employed and were not vaccinated at baseline (June 2021) but received vaccination at a four-month follow-up (October 2021). An 11-item scale measured the side effects of COVID-19 vaccines. Four types of psychosocial working conditions (i.e., job demands, job control, and supervisor and coworker support) were measured using the Brief Job Stress Questionnaire (BJSQ). Multiple linear regression analyses were conducted to examine the relationship between the psychosocial working conditions and side effects of COVID-19 vaccines, adjusting for gender, age, educational attainment, marital status, occupation, chronic disease, dose of vaccination, anxiety from potential side effects of vaccines, fear and worry about COVID-19, and psychological distress at baseline.

**Results:** Overall, 747 employees were included in the analysis. The average number of side effects was 3.78 (SD=2.19): Arm pain (81.1%), fatigues (64.1%), muscle pains (63.3%), and fever (37.5 degrees Celsius +) (53.5%) were reported more frequently. Coworker support score was significantly and negatively associated with the numbers of side effects (standardized β=-0.122, p=0.017). Women, young age, second time vaccination, and high psychological distress were significantly associated with several side effects.

**Conclusions:** Employees with low coworker support may be more likely to have side effects after COVID-19 vaccinations. The findings of this study could inform employees with low coworker support that increasing workplace support may reduce the side effects.

**Highlights:**

⍰ The effect of poor psychosocial working conditions on side effects after COVID-19 vaccinations was unknown.
⍰ Poor coworker support at baseline was significantly associated with increased side effects after COVID-19 vaccinations.
⍰ Informing workers with low coworker support about the findings may help them prepare for the side effect and motivate them to improve coworker support to reduce side effects.

## Introduction

Solicited local and systemic adverse events, that is, side effects after the injection of COVID-19 vaccines, have been frequently reported [1, 2]. They affect the daily life activities of the recipients. They may also be a major reason for vaccine hesitancy [3]. The immediate and non-specific innate immune response can produce various side effects [4]. Women, young people, second dose, heterologous prime-boost, and individuals with previous SARS-CoV-2 infection are more likely to experience side effects of the COVID-19 vaccine [5, 6]. Differences in side effects have been attributed to increased immunogenicity to the COVID-19 vaccine among these groups [5,7,8].

Psychological factors can affect the immune system’s response to the vaccine, thus the side effect [4]. After influenza virus vaccination, for example, chronic depression was associated with excessive and prolonged inflammatory responses [7]. Exposures to a brief stressor before receiving the typhoid vaccine amplified the inflammatory response to the vaccine [8]. Psychological factors may also trigger short-term side effects of COVID-19 vaccination. However, to date, no study has examined this association.

Poor psychosocial working conditions, such as high job demands, low job control [9, 10], and lack of workplace support [11], have been associated with immune system dysregulation [12-15]. Poor psychosocial working conditions often increase inflammatory markers while reducing cellular immune functions (such as NK cell activity, NK and T cell subsets, CD4+/CD8+ ratio) [16]. Thus, people working under poor psychosocial conditions may experience more side effects after the COVID-19 vaccine because of decreased innate immune response to vaccination caused by such stressful conditions.

This prospective study aimed to examine the association of job demands, job control, and lack of workplace support with side effects after receiving COVID-19 vaccination in a sample of full-time employees in Japan.

## Methods

### Study design and participants

The data were collected as a part of a large-scale prospective panel study, the Employee Cohort Study in the Covid-19 pandemic in Japan (E- COCO- J) [17, 18], targeting a sample of full-time employees recruited from the panel of the Japanese internet company in March 2020 (N=1448). After completing six surveys (including the first survey) between March 2020 and March 2021, the seventh and eighth surveys were administered in June and October 2021, respectively. In this prospective study, the baseline variables (such as job strain and workplace support) were measured in the seventh survey (hereafter called the baseline), and the outcome variables (i.e., the side effect of the COVID-19 vaccine) were measured in the eighth survey (hereafter called as the follow-up). The participants’ eligibility criteria were: 1) being employed at baseline, 2) not being vaccinated at baseline, and 3) being vaccinated at follow-up. The details of the recruitment process are shown in **Figure 1**.

The Research Ethics Committee of the Graduate School of Medicine/Faculty of Medicine, The University of Tokyo, approved this study, No. 10856-(2)(3)(4)(5).

### Measurement variables

#### Side effects of COVID-19 vaccine

A list of 11 side effects of the COVID-19 vaccine was created by referring to the report of the possible common side effects reported by the Centers for Disease Control and Prevention (CDC) [19], including arm pain/redness/swelling, fatigues/tiredness, headache, muscle pain/joint pain, chills, fevers (37.5 degree+), nausea/vomit, diarrhea, lymph node pain, severe reactions to being needed medical care (e.g., anaphylaxes), and delayed local arm reactions after 7 days of vaccinations (i.e., COVID arm). We developed an 11-item scale of side effects of COVID-19 vaccines. Participants were asked whether they had each of the listed side effects within a few days after the vaccination: “Did you experience any of the following side effects within 1 to 3 days after getting a COVID-19 vaccine?” If respondents received only the first dose, they were asked to report their experience at that time; if they received the vaccination twice, they were asked to report their experience when their symptoms were most severe. The response options were Yes or No. The total number of side effects, ranging from 0 to 11, was used as a primary outcome.

#### Psychosocial working conditions

Psychosocial working conditions were assessed based on the job-demand-control (JDC) model and the job-demand-control-support (JDCS) model, which explain the occurrence of mental strain in a workplace context [9, 20]. Four components of the JDC/JDCS model, job demand (quantitative job overload), job control, coworker support, and supervisor support, were measured using the corresponding subscales of the Brief Job Stress Questionnaire (BJSQ) [21]. Each scale comprised three items, with each being rated on a 4-point Likert-type scale from ‘very much so’ = 1 to ‘not at all’ = 4 for job demands and job control and from ‘Extremely’ = 4 to ‘not at all’ = 1) for supervisor and coworker support. Total scores for each subscale ranged from 3 to 12, with a higher score indicating the higher degree of the corresponding component. The four components of the BJSQ showed good reliability and validity.

#### Covariates

##### Anxiety about side effects of vaccines

One original item was used to assess anxiety about the side effects of vaccines. This item stated, “I am concerned about the effectiveness and side effects of COVID-19 vaccines,” Responses were scored on a 4-point Likert-type scale (ranging from 1 “Not at all” to 4 “Feel strong”). The reliability and validity were not examined.

##### Fear and worry about COVID-19

One original item was used to assess participants’ fear of COVID-19. The item asked, “Do you feel anxious about COVID-19?” Responses were scored on a 6-point Likert-type scale (ranging from 1 “Not at all” to 6 “Feel strongly”). The reliability and validity were not examined, but several papers using the same scale were published [17, 22, 23].

##### Psychological distress

Psychological distress was measured using K6 (Kessler 6) [24, 25]. Respondents were asked to report how frequently in the past 4 weeks they felt nervous, hopeless, restless or fidgety, worthless, depressed, and that everything was an effort. The response options were “none of the time”=0, “a little of the time”=1, “some of the time”=2, “most of the time”=3, or “all of the time”=4. The total score ranges from 0 to 24. The Japanese version of K6 showed good reliability and validity [26]. K6 performs just as well as the Composite International Diagnostic Interview (CIDI) Short Form in identifying individuals with clinically significant mental disorders [27].

##### Demographic variables

We measured gender, age, educational attainment, marital status, occupation, chronic disease at baseline, and dose of vaccination at follow-up. Chronic disease was defined as having any physical and psychological diseases which were currently treated in the medical settings, including hypertension, diabetes, heart disease (e.g., angina, heart failure), cerebrovascular disease (e.g., cerebral infarction, cerebral hemorrhage), cancer/malignant neoplasm, respiratory disease, liver disease, kidney disease, and depression/anxiety/unstable moods. Hospital admissions or home treatment over one week regardless of COVID-19 in the past 6 months was measured at the follow-up.

### Statistical analysis

The descriptive statistics were used to describe sociodemographic characteristics, psychosocial working conditions, and the prevalence of reported side effects. We conducted multiple linear regression analyses to examine the relationship between four psychosocial factors at work (job demand, job control, coworker’s support, and supervisor support) and the side effects of COVID-19 vaccines in the crude model (Model 1). The relationship was also assessed when adjusting for gender, age, educational attainment, marital status, occupation, chronic disease, and dose of vaccination (Model 2) and for anxiety about side effects of vaccines, fear and worry about COVID-19 and psychological distress at baseline (Model 3). The same subgroup analysis was conducted among participants who received second vaccination.

The sample may have included participants who tested positive for COVID-19 before the baseline or during the follow-up, which may have confounded the results. We were not allowed to ask whether the participants were infected in the survey for ethical reasons. Instead, we asked whether they received any in-hospital or home treatment for one week or longer in the past six months to exclude respondents who were potentially infected. As a sensitivity analysis, we conducted the same analysis excluding participants who reported hospital admissions or home treatment for one week or longer during the past 6 months. The primary outcome was the total number of side effects.

Multiple logistic regression analysis was conducted to examine the relationship between four psychosocial factors at work and severe adverse effects requiring medical care after getting vaccinated (i.e., anaphylaxis). SPSS 28.0 (IBM Corp) Japanese version was used. Statistical significance was set as a two-sided p < .05.

## Results

The participants’ characteristics are shown in **Table 1**. The mean age was 44.8 years old (min-max: 22-62 years old). Those with chronic disease accounted for 14.5%, and 8.6% of the participants experienced hospital admissions or home treatment over one week regardless of COVID-19 during the past 6 months.

**Table 1.** Participants’ sociodemographic characteristics and psychosocial factors at work at baseline (N=747).

| Variables [possible range] | N (%) | Mean (SD) |
| --- | --- | --- |
| Age |  | 44.79 (10.18) |
| 20 – 29 years old | 62 (8.3) |  |
| 30 – 39 years old | 173 (23.2) |  |
| 40 – 49 years old | 233 (31.2) |  |
| 50+ | 279 (37.3) |  |
| Gender |  |  |
| Men | 423 (56.6) |  |
| Women | 324 (43.4) |  |
| Marital status |  |  |
| Single | 320 (42.8) |  |
| Married | 427 (57.2) |  |
| Educational attainment (a) |  |  |
| Less than a high school diploma | 145 (19.3) |  |
| College/vocational | 16.2 (16.2) |  |
| Undergraduate | 296 (39.6) |  |
| Graduate+ | 37 (5.0) |  |
| Unknown/others | 148 (19.8) |  |
| Occupation (b) |  |  |
| Mangers | 93 (12.4) |  |
| Non-manual workers | 447 (59.8) |  |
| Manual workers | 184 (24.6) |  |
| Healthcare workers | 23 (3.1) |  |
| Chronic disease (c) |  |  |
| None | 639 (85.5) |  |
| Any | 108 (14.5) |  |
| Hospital admissions/ home treatment (d) |  |  |
| No | 683 (91.4) |  |
| Yes | 64 (8.6) |  |
| Vaccination |  |  |
| First shot | 60 (8.0) |  |
| Second shot | 687 (92.0) |  |
| Anxiety about side effects of vaccination [1-4] |  | 2.99 (0.82) |
| Fear and worry about COVID-19 [1-6] |  | 4.26 (1.23) |
| Job demand [3-12] |  | 7.92 (2.35) |
| Job control [3-12] |  | 8.00 (2.19) |
| Coworker support [3-12] |  | 6.95 (2.15) |
| Supervisor support [3-12] |  | 6.65 (2.21) |
| Psychological distress [0-24] |  | 5.55 (5.73) |

SD: Standard deviation.
(a) Information about educational attainment was obtained in May 2020.
(b) Information about the occupation was created by merging the data obtained in February 2019 with information of the job category (i.e., healthcare workers or not) in May 2020.
(d) Hospital admissions or home treatment over one week, regardless of COVID-19, in the past 6 months.

The prevalence rates of self-reported side effects after getting a COVID-19 vaccine are shown in **Table 2**. The most prevalent side effects were arm pain/redness/swelling (81.1%), fatigues/tiredness (64.1%), muscle pains/joint pains (63.3%), and fever (37.5 degree Celsius +) (53.5%). In contrast, severe reactions requiring needed medical care (e.g., anaphylaxes) accounted for 2.9%.

**Table 2.**
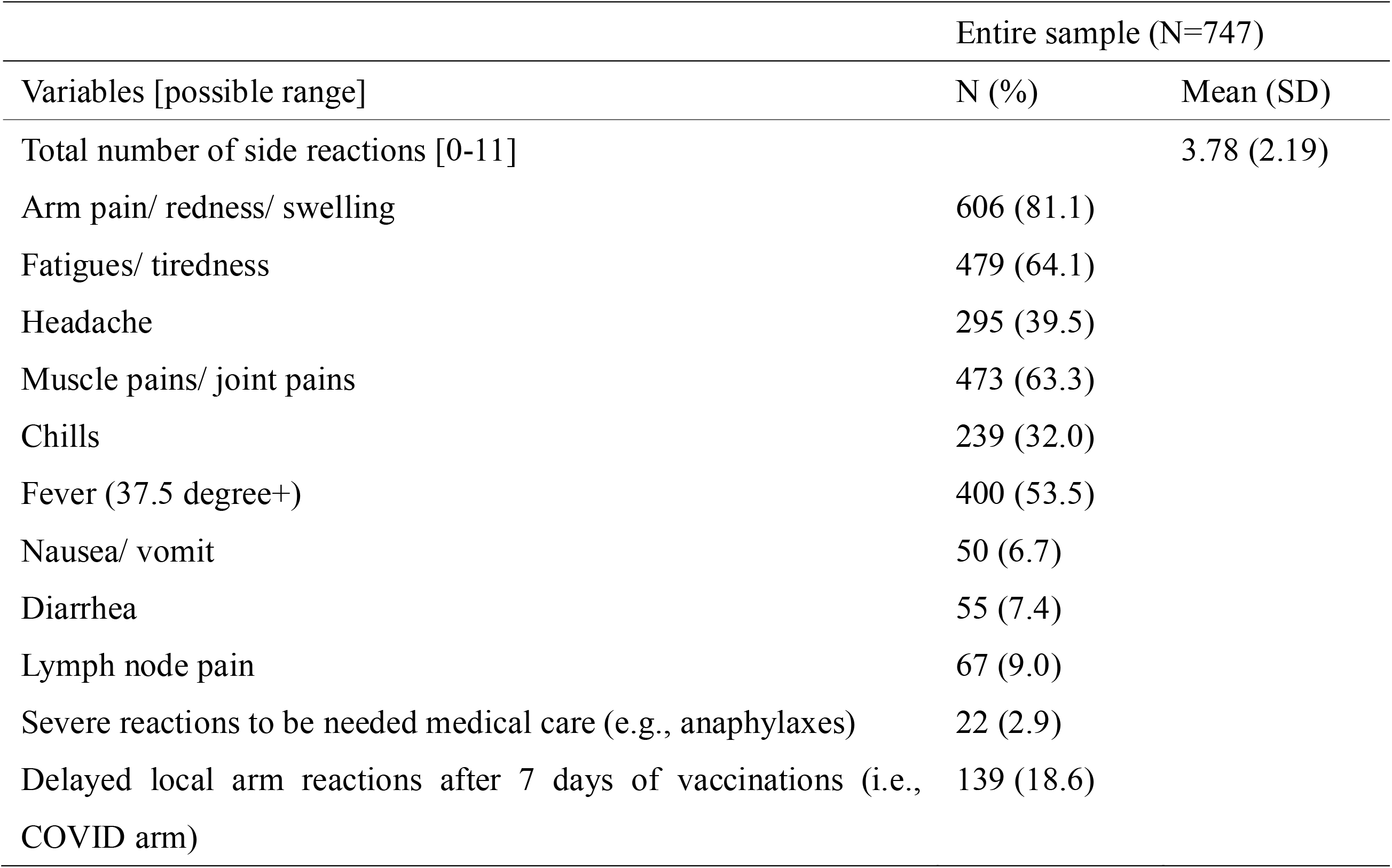
Prevalence of self-reported side effects after getting a COVID-19 vaccine.

The results of multiple linear regression analysis are shown in **Table 3**. Job demand and coworker support were significantly associated with the number of side effects in Model 2 (standardized β=0.081, p=0.041; β=-0.134, p=0.008, respectively). Coworker support also showed significance in Model 3 (β=-0.122, p=0.017). Psychological distress at baseline was significantly associated with side effects in Model 3 (β=0.150, p<0.001). Women, younger age, and second vaccination were significantly associated with the great number of the side effects.

**Table 3.**
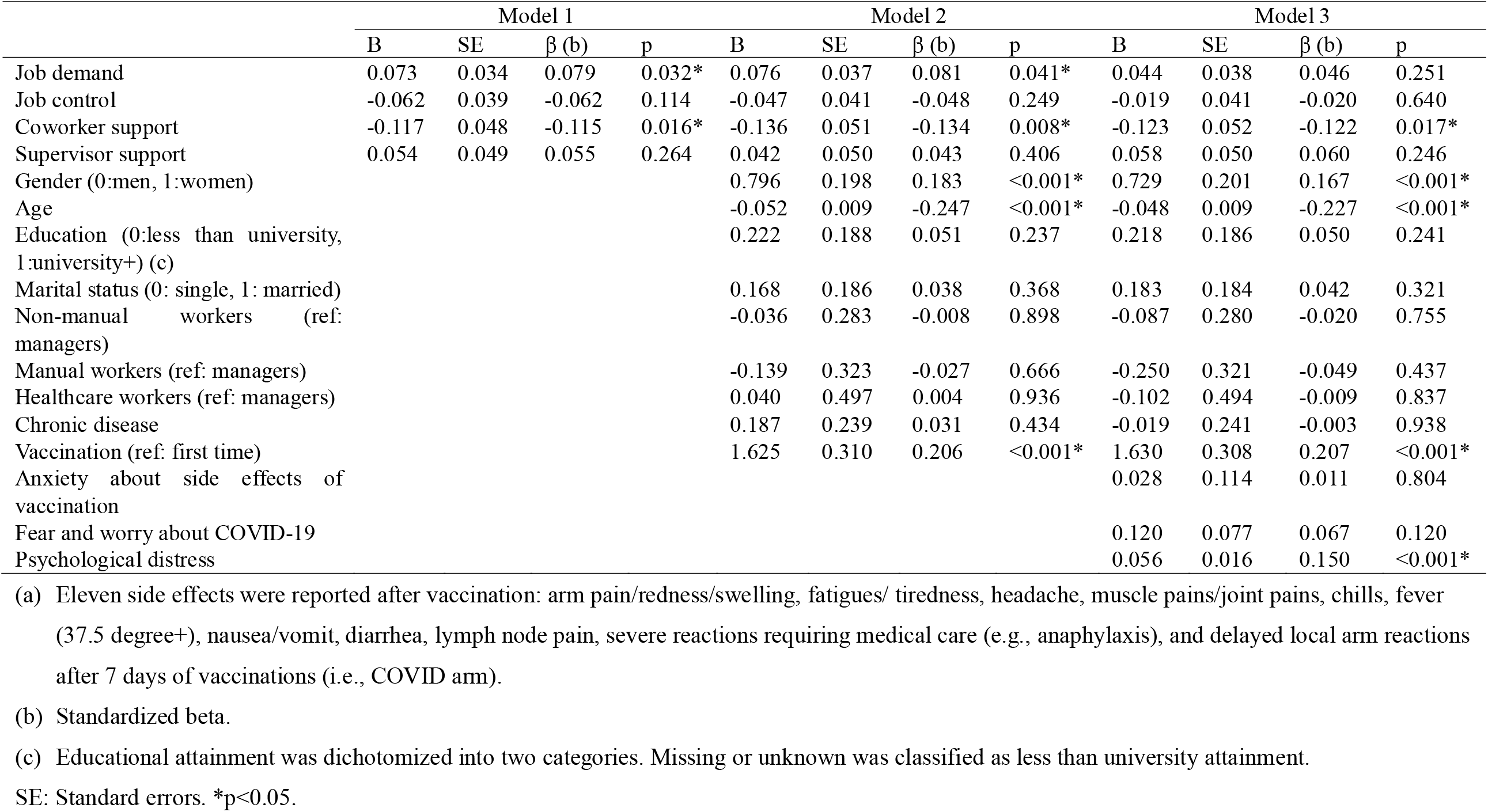
Association between psychosocial factors at work and the total number of side effects (a) of COVID-19 vaccines in the entire sample (N=747): Multiple linear regression analysis.

The subgroup analysis with participants who received second vaccination (N=687) showed the negative associations of coworker support with the total number of side effects in Model 3 (β=-0.115, p=0.034) (**Appendix 1**).

The result of multiple linear regression analysis among those who did not experience hospital admissions or home treatment during the past six months (N=683) is shown in **Appendix 2**. In Model 3 of the multiple linear regression analysis (**Appendix 3**), coworker support was significantly negatively associated with the number of side effects (β=-0.131, p=0.014).

None of the four psychosocial working conditions correlated significantly with the presence of severe adverse reactions (1 item) in the adjusted model in the entire sample (N=747) (**Appendix 4A**) or among participants without hospital admissions/home treatment (N=683) (**Appendix 4B**).

## Discussion

High coworker support was positively associated with fewer side effects after the COVID-19 vaccinations. Job demand, control, and supervisor support did not show significant associations. However, high psychological distress was associated with side effects. This study demonstrated the importance of psychosocial working conditions in employees experiencing various side effects after the COVID-19 vaccinations.

Coworker support showed a significant negative association with the total number of side effects of the COVID-19 vaccine. The finding is consistent with previous findings showing that psychological factors, i.e., exposure to a stressor, precipitate immediate inflammatory reactions to vaccines [4, 7] and that poor psychosocial working conditions often increase inflammatory markers [16]. However, this association was observed even after adjusting for baseline psychological distress, fear, and worry about COVID-19. Although psychological distress may partially mediate the association, poor coworker support may be independently associated with the side effect of the COVID-19 vaccine. The possible mechanism underlying the association is unclear, but the finding that the lack of social support generally increases systematic inflammations might clarify this mechanism [4, 12, 28, 29]. Mediators and products that cause inflammation in the circulation can affect body systems to cause systemic side effects [30]. For instance, a previous study indicated that low coworker support influenced inflammation biomarkers in a group of employees who worked more than 41h per week [12]. It may be plausible that elevated inflammatory responses associated with lack of coworker support may exaggerate an innate immune response to the COVID-19 vaccine [31, 32].

Job demand, control, or supervisor support showed no significant associations with reported side effects. However, these associations may have been underestimated due to this study’s relatively small sample size. These psychosocial working conditions are also related to chronic inflammations; for example, previous studies have indicated that high supervisor support was associated with low inflammation markers [12, 33]. Hence, environmental factors may potentially affect the experience of side effects. Future investigations should use larger samples to replicate the findings.

### Strengths and limitations

The strength of this study was the prospective nature of the study design. This study showed direct and indirect effects of the psychosocial working conditions on the immune function. However, the present study had several limitations. First, self-reported measures were used to assess the psychosocial working conditions and side effects. People with poor working conditions or high distress may have overreported the side effects. Besides, employees with low coworker support may be easily conscious of side effects because fewer people around them take over their work, so they are more concerned about the impact on their work. Second, this study did not consider other psychosocial conditions other than job demand, job control, and workplace social support. Other factors should be investigated in future research. Third, generalizability was limited because the participants were full-time employees in Japan, and the data were retrieved from the online panel. Fourth, the significant association found in this study could have been superficial, as other potential confounding factors could have affected the results. Fifth, this study addressed only a very short-term innate immune response to vaccination but did not examine the effects on cellular or humoral immunity after vaccination and over longer period. The previous meta-analysis revealed a negative association between stress and antibody production after influenza vaccinations [34]. In addition, several studies have suggested that lonely or socially isolated individuals had a weak immune response after vaccinations [4, 35, 36] and increased susceptibility to infectious disease [37]. Future studies can examine the association of psychosocial working conditions and psychological stress with decreased responses to vaccinations and infectious susceptibility.

### Implications for future practice

This study demonstrated the importance of psychosocial working conditions, especially coworker support, in experiencing certain adverse events after the COVID-19 vaccinations. Informing employees about the potential effects of low coworker support on adverse vaccine outcomes may help them prepare for these side effects. Improving coworker support in the workplace may reduce some of the reported side effects. Active actions to control the reactogenicity may ease the vaccine hesitancy. Even before COVID-19, stressors and stress reactions have been known to be important factors in immune functions. Accordingly, they should receive even greater attention during the pandemic. Further studies are needed to examine the association of social supports with immune responses.

## Conclusions

Coworker support showed a significantly negative association with experiencing short-term side effects. Providing information about the findings may help employees cope and prepare for potential side effects. Future research is needed to replicate the findings with large sample size.

## Data Availability

The data supporting this study's findings are available from the corresponding author, NK, upon reasonable request.

## Acknowledgment

We appreciate the involvement of all E-COCO-J study participants and collaborators.

## References

1. Chapin-Bardales, J., J. Gee, and T. Myers, Reactogenicity Following Receipt of mRNA-Based COVID-19 Vaccines. JAMA, 2021. 325(21): p. 2201–2202.

2. Baden, L.R., et al., Efficacy and Safety of the mRNA-1273 SARS-CoV-2 Vaccine. N Engl J Med, 2021. 384(5): p. 403–416.

3. Biswas, M.R., et al., A Scoping Review to Find Out Worldwide COVID-19 Vaccine Hesitancy and Its Underlying Determinants. Vaccines (Basel), 2021. 9(11).

4. Madison, A.A., et al., Psychological and Behavioral Predictors of Vaccine Efficacy: Considerations for COVID-19. Perspect Psychol Sci, 2021. 16(2): p. 191–203.

5. Menni, C., et al., Vaccine side-effects and SARS-CoV-2 infection after vaccination in users of the COVID Symptom Study app in the UK: a prospective observational study. Lancet Infect Dis, 2021. 21(7): p. 939–949.

6. Saita, M., et al., Reactogenicity following two doses of the BNT162b2 mRNA COVID-19 vaccine: Real-world evidence from healthcare workers in Japan. Journal of infection and chemotherapy: official journal of the Japan Society of Chemotherapy, 2022. 28(1): p. 116–119.

7. Glaser, R. et al., Mild depressive symptoms are associated with amplified and prolonged inflammatory responses after influenza virus vaccination in older adults. Archives of general psychiatry, 2003. 60(10): p. 1009–1014.

8. Brydon, L., et al., Synergistic effects of psychological and immune stressors on inflammatory cytokine and sickness responses in humans. Brain, behavior, and immunity, 2009. 23(2): p. 217–224.

9. Karasek Jr, R.A., Job demands, job decision latitude, and mental strain: Implications for job redesign. Administrative science quarterly, 1979: p. 285–308.

10. Karasek, R., & Theorell, T, Healthy Work: Stress, Productivity, and the Reconstruction of Working Life. 1990, New York: Basic Books.

11. Johnson, J.V. and E.M. Hall, Job strain, workplace social support, and cardiovascular disease: a cross-sectional study of a random sample of the Swedish working population. Am J Public Health, 1988. 78(10): p. 1336–42.

12. Eguchi, H., et al., Source - specific workplace social support and high - sensitivity C - reactive protein levels among Japanese workers: A 1 - year prospective cohort study. American journal of industrial medicine, 2016. 59(8): p. 676–684.

13. Shirom, A., et al., The Job Demand-Control-Support Model and stress-related low-grade inflammatory responses among healthy employees: A longitudinal study. Work & Stress, 2008. 22(2): p. 138–152.

14. Nabi, H. et al., Do psychological factors affect inflammation and incident coronary heart disease: the Whitehall II Study. Arteriosclerosis, thrombosis, and vascular biology, 2008. 28(7): p. 1398–1406.

15. Sara, J.D., et al., Association Between Work&Related Stress and Coronary Heart Disease: A Review of Prospective Studies Through the Job Strain, Effort&Reward Balance, and Organizational Justice Models. Journal of the American Heart Association, 2018. 7(9): p. e008073.

16. Nakata, A., Psychosocial job stress and immunity: a systematic review. Methods Mol Biol, 2012. 934: p. 39–75.

17. Kawakami, N. et al., The Effects of Downloading a Government-Issued COVID-19 Contact Tracing App on Psychological Distress During the Pandemic Among Employed Adults: Prospective Study. JMIR Ment Health, 2021. 8(1): p. e23699.

18. Sasaki, N., et al., The deterioration of mental health among healthcare workers during the COVID-19 outbreak: A population-based cohort study of workers in Japan. Scand J Work Environ Health, 2020. 46(6): p. 639–644.

19. (CDC);, C.f.D.C.a.P. Possible Side Effects After Getting a COVID-19 Vaccine. 2021 [cited 2021 November 18]; Available from: https://www.cdc.gov/coronavirus/2019-ncov/vaccines/expect/after.html.

20. Johnson, J.V., E.M. Hall, and T. Theorell, Combined effects of job strain and social isolation on cardiovascular disease morbidity and mortality in a random sample of the Swedish male working population. Scand J Work Environ Health, 1989. 15(4): p. 271–9.

21. Shimomitsu, T., et al., Investigation research report concerning prevention of disease related to work in 1997 the Ministry of Labor: III Stress measurement research group report. Tokyo: Tokyo Medical University, 2000: p. 101–169.

22. Sasaki, N., et al., Workplace responses to COVID-19 associated with mental health and work performance of employees in Japan. J Occup Health, 2020. 62(1): p. e12134.

23. Sasaki, N., et al., Exposure to media and fear and worry about COVID-19. Psychiatry Clin Neurosci, 2020. 74(9): p. 501–502.

24. Kessler, R.C., et al., Short screening scales to monitor population prevalences and trends in non-specific psychological distress. Psychol Med, 2002. 32(6): p. 959–76.

25. Kessler, R.C., et al., Screening for serious mental illness in the general population. Arch Gen Psychiatry, 2003. 60(2): p. 184–9.

26. Sakurai, K., et al., Screening performance of K6/K10 and other screening instruments for mood and anxiety disorders in Japan. Psychiatry Clin Neurosci, 2011. 65(5): p. 434–41.

27. Kessler, R.C., et al., Screening for Serious Mental Illness in the General Population. Archives of General Psychiatry, 2003. 60(2): p. 184–189.

28. Andrew P S., The Psychology of the Common Cold and Influenza: Implications for COVID-19. International Journal of Clinical Virology, 2020. 4(1): p. 027–031.

29. Uchino, B.N., et al., Social support, social integration, and inflammatory cytokines: A meta-analysis. Health Psychology, 2018. 37(5): p. 462.

30. Herve, C., et al., The how’s and what’s of vaccine reactogenicity. NPJ Vaccines, 2019. 4: p. 39.

31. Mortensen, R.F., C-reactive protein, inflammation, and innate immunity. Immunol Res, 2001. 24(2): p. 163–76.

32. Tanaka, T., M. Narazaki, and T. Kishimoto, IL-6 in inflammation, immunity, and disease. Cold Spring Harb Perspect Biol, 2014. 6(10): p. a016295.

33. Nakata, A., M. Irie, and M. Takahashi, Source-Specific Social Support and Circulating Inflammatory Markers Among White-Collar Employees. Annals of Behavioral Medicine, 2013. 47(3): p. 335–346.

34. Pedersen, A.F., R. Zachariae, and D.H. Bovbjerg, Psychological stress and antibody response to influenza vaccination: a meta-analysis. Brain Behav Immun, 2009. 23(4): p. 427–33.

35. Glaser, R., et al., Stress-induced modulation of the immune response to recombinant hepatitis B vaccine. Psychosomatic medicine, 1992. 54(1): p. 22–29.

36. Gallagher, S. et al., Psychosocial factors are associated with the antibody response to both thymus-dependent and thymus-independent vaccines. Brain, behavior, and immunity, 2008. 22(4): p. 456–460.

37. Cohen, S., et al., Social ties and susceptibility to the common cold. Jama, 1997. 277(24): p. 1940–1944.

