## Appendix for "The effect of job strain and worksite social support on reported side effects of COVID-19 vaccine: a prospective study of employees in Japan"

**Appendix 1. Association between psychosocial factors at work and the total number of side effects (a) after the second COVID-19 vaccination (N=687): Multiple linear regression analysis.**

|  | Model 1 | | | | | Model 2 | | | | Model 3 | | | |
| --- | --- | --- | --- | --- | --- | --- | --- | --- | --- | --- | --- | --- | --- |
|  | B | SE | β (b) | p | B | | SE | β (b) | p | B | SE | β (b) | p |
| Job demand | 0.071 | 0.036 | 0.076 | 0.048 | 0.081 | | 0.040 | 0.085 | 0.041 | 0.048 | 0.040 | 0.051 | 0.231 |
| Job control | -0.075 | 0.041 | -0.075 | 0.069 | -0.056 | | 0.043 | -0.057 | 0.202 | -0.03 | 0.043 | -0.031 | 0.484 |
| Coworker support | -0.099 | 0.051 | -0.096 | 0.053 | -0.126 | | 0.054 | -0.123 | 0.021 | -0.117 | 0.055 | -0.115 | 0.034* |
| Supervisor support | 0.048 | 0.051 | 0.048 | 0.351 | 0.030 | | 0.054 | 0.031 | 0.572 | 0.054 | 0.054 | 0.055 | 0.315 |
| Gender (0: men, 1: women) |  |  |  |  | 0.896 | | 0.212 | 0.205 | <.001* | 0.823 | 0.214 | 0.188 | <.001* |
| Age |  |  |  |  | -0.055 | | 0.010 | -0.259 | <.001* | -0.05 | 0.010 | -0.236 | <.001* |
| Education (0: less than university, 1: university+) (c) |  |  |  |  | 0.257 | | 0.200 | 0.059 | 0.200 | 0.256 | 0.198 | 0.059 | 0.197 |
| Marital status (0: single, 1: married) |  |  |  |  | 0.212 | | 0.200 | 0.048 | 0.289 | 0.221 | 0.198 | 0.050 | 0.264 |
| Non-manual workers (ref: managers) |  |  |  |  | -0.103 | | 0.295 | -0.023 | 0.728 | -0.162 | 0.294 | -0.037 | 0.581 |
| Manual workers (ref: managers) |  |  |  |  | -0.032 | | 0.340 | -0.006 | 0.925 | -0.155 | 0.339 | -0.030 | 0.646 |
| Healthcare workers (ref: managers) |  |  |  |  | 0.114 | | 0.547 | 0.010 | 0.835 | -0.017 | 0.542 | -0.001 | 0.974 |
| Chronic disease |  |  |  |  | 0.161 | | 0.251 | 0.027 | 0.521 | -0.062 | 0.255 | -0.010 | 0.807 |
| Anxiety about side effects of vaccination |  |  |  |  |  | |  |  |  | 0.064 | 0.122 | 0.024 | 0.599 |
| Fear and worry about COVID-19 |  |  |  |  |  | |  |  |  | 0.093 | 0.082 | 0.052 | 0.255 |
| Psychological distress |  |  |  |  |  | |  |  |  | 0.059 | 0.017 | 0.154 | <.001* |

(a) Eleven side effects were reported after vaccination: arm pain/redness/swelling, fatigues/ tiredness, headache, muscle pains/joint pains, chills, fever (37.5 degree+), nausea/vomit, diarrhea, lymph node pain, severe reactions requiring medical care (e.g., anaphylaxis), and delayed local arm reactions after 7 days of vaccinations (i.e., COVID arm).
(b) Standardized beta.
(c) Educational attainment was dichotomized into two categories. Missing or unknown was classified as less than university attainment.

SE: Standard errors. *p<0.05.

**Appendix 2. Prevalence of self-reported side effects after getting a COVID-19 vaccine among the participants who experienced hospital admissions or home treatment in the last 6 months lasting over one week (N=64).**

|  | Hospital admissions/home treatment (N=64) | |
| --- | --- | --- |
| Variables [possible range] | N (%) | Mean (SD) |
| Total number of side reactions [0-11] |  | 3.98 (2.26) |
| Arm pain/ redness/ swelling | 55 (85.9) |  |
| Fatigues/ tiredness | 44 (68.8) |  |
| Headache | 24 (37.5) |  |
| Muscle pains/ joint pains | 38 (59.4) |  |
| Chills | 21 (32.8) |  |
| Fever (37.5 degree+) | 34 (53.1) |  |
| Nausea/ vomit | 5 (7.8) |  |
| Diarrhea | 6 (9.4) |  |
| Lymph node pain | 7 (10.9) |  |
| Severe reactions to be needed medical care (e.g., anaphylaxis) | 6 (9.4) |  |
| Delayed local arm reactions after 7 days of vaccinations (i.e., COVID arm) | 15 (23.4) |  |

**Appendix 3. Association between psychosocial factors at work and the total number of side effects (a) after COVID-19 vaccination among participants without hospital admissions/home treatment (N=683): Multiple linear regression analysis.**

|  | Model 1 | | | | | Model 2 | | | | Model 3 | | | |
| --- | --- | --- | --- | --- | --- | --- | --- | --- | --- | --- | --- | --- | --- |
|  | B | SE | β (b) | p | B | | SE | β (b) | p | B | SE | β (b) | p |
| Job demand | 0.074 | 0.035 | 0.081 | 0.034* | 0.075 | | 0.038 | 0.081 | 0.050 | 0.041 | 0.039 | 0.044 | 0.296 |
| Job control | -0.076 | 0.041 | -0.078 | 0.060 | -0.064 | | 0.042 | -0.067 | 0.130 | -0.032 | 0.042 | -0.034 | 0.444 |
| Coworker support | -0.120 | 0.051 | -0.118 | 0.019* | -0.140 | | 0.053 | -0.138 | 0.009* | -0.132 | 0.054 | -0.131 | 0.014* |
| Supervisor support | 0.069 | 0.051 | 0.070 | 0.177 | 0.064 | | 0.053 | 0.066 | 0.228 | 0.083 | 0.053 | 0.085 | 0.116 |
| Gender (0: men, 1: women) |  |  |  |  | 0.765 | | 0.207 | 0.176 | <0.001* | 0.696 | 0.208 | 0.160 | <0.001* |
| Age |  |  |  |  | -0.054 | | 0.010 | -0.257 | <0.001* | -0.051 | 0.010 | -0.240 | <0.001* |
| Education (0: less than university, 1: university+) (c) |  |  |  |  | 0.219 | | 0.195 | 0.050 | 0.264 | 0.206 | 0.193 | 0.048 | 0.286 |
| Marital status (0: single, 1: married) |  |  |  |  | 0.122 | | 0.194 | 0.028 | 0.528 | 0.135 | 0.191 | 0.031 | 0.479 |
| Non-manual workers (ref: managers) |  |  |  |  | -0.139 | | 0.295 | -0.032 | 0.637 | -0.174 | 0.291 | -0.040 | 0.550 |
| Manual workers (ref: managers) |  |  |  |  | -0.225 | | 0.334 | -0.044 | 0.502 | -0.340 | 0.331 | -0.067 | 0.305 |
| Healthcare workers (ref: managers) |  |  |  |  | 0.282 | | 0.564 | 0.023 | 0.618 | 0.160 | 0.558 | 0.013 | 0.775 |
| Chronic disease |  |  |  |  | 0.153 | | 0.250 | 0.025 | 0.540 | -0.060 | 0.252 | -0.010 | 0.812 |
| Vaccination (ref: first time) |  |  |  |  | 1.527 | | 0.345 | 0.181 | <0.001* | 1.480 | 0.342 | 0.176 | <0.001* |
| Anxiety about side effects of vaccination |  |  |  |  |  | |  |  |  | 0.007 | 0.119 | 0.003 | 0.956 |
| Fear and worry about COVID-19 |  |  |  |  |  | |  |  |  | 0.162 | 0.080 | 0.092 | 0.042* |
| Psychological distress |  |  |  |  |  | |  |  |  | 0.057 | 0.017 | 0.150 | <0.001* |

(d) Eleven side effects were observed after vaccination: arm pain/redness/swelling, fatigues/ tiredness, headache, muscle pains/joint pains, chills, fever (37.5 degree+), nausea/vomit, diarrhea, lymph node pain, severe reactions requiring medical care (e.g., anaphylaxis), and delayed local arm reactions after 7 days of vaccinations (i.e., COVID arm).
(e) Standardized beta.
(f) Educational attainment was dichotomized into two categories. Missing or unknown was classified as less than university attainment.

SE: Standard errors. *p<0.05.

**Appendix 4A. Association between psychosocial factors at work and severe adverse effects (a) of COVID-19 vaccines: Multiple logistic regression analysis** **in the entire sample (N=747).**

|  | **Model 1** | | | **Model 2** | | | **Model 3** | | |
| --- | --- | --- | --- | --- | --- | --- | --- | --- | --- |
|  | OR | 95% CI | p | OR | 95% CI | p | OR | 95% CI | p |
| Job demand | 0.98 | 0.81 - 1.18 | 0.814 | 0.99 | 0.79 - 1.25 | 0.945 | 1.06 | 0.83 - 1.36 | 0.629 |
| Job control | 0.97 | 0.78 - 1.19 | 0.748 | 1.00 | 0.78 - 1.28 | 0.982 | 0.99 | 0.77 - 1.28 | 0.961 |
| Coworker support | 0.79 | 0.61 - 1.02 | 0.074 | 0.82 | 0.6 - 1.12 | 0.207 | 0.88 | 0.63 - 1.22 | 0.433 |
| Supervisor support | 1.37 | 1.06 - 1.77 | 0.015* | 1.36 | 1 - 1.86 | 0.050 | 1.31 | 0.96 - 1.8 | 0.092 |
| Gender (0: men, 1: women) |  |  |  | 2.34 | 0.7 - 7.81 | 0.168 | 3.48 | 0.94 - 12.92 | 0.062 |
| Age |  |  |  | 0.98 | 0.92 - 1.03 | 0.394 | 0.98 | 0.92 - 1.04 | 0.446 |
| Education (0: less than university, 1: university+) (b) |  |  |  | 0.63 | 0.2 - 1.98 | 0.425 | 0.65 | 0.2 - 2.08 | 0.469 |
| Marital status (0: single, 1: married) |  |  |  | 2.13 | 0.69 - 6.63 | 0.191 | 2.43 | 0.77 - 7.72 | 0.132 |
| Non-manual workers (ref: managers) |  |  |  | 1.42 | 0.15 - 13.74 | 0.764 | 1.73 | 0.18 - 16.8 | 0.638 |
| Manual workers (ref: managers) |  |  |  | 1.65 | 0.15 - 18.61 | 0.686 | 2.15 | 0.19 - 24.97 | 0.541 |
| Healthcare workers (ref: managers) |  |  |  | 3.03 | 0.15 - 59.81 | 0.467 | 3.03 | 0.15 - 61.12 | 0.469 |
| Chronic disease (ref: managers) |  |  |  | 0.45 | 0.06 - 3.68 | 0.459 | 0.57 | 0.07 - 4.77 | 0.601 |
| Vaccination (ref: first time) |  |  |  | 47252283.19 |  | 0.997 | 46938379.70 |  | 0.997 |
| Anxiety about side effects of vaccination |  |  |  |  |  |  | 0.74 | 0.37 - 1.47 | 0.387 |
| Fear and worry about COVID-19 |  |  |  |  |  |  | 0.70 | 0.44 - 1.12 | 0.137 |
| Psychological distress |  |  |  |  |  |  | 1.01 | 0.92 - 1.12 | 0.829 |

(a) Severe adverse effect was defined as self-reported severe reactions requiring medical care after vaccination (e.g., anaphylaxis). Severe adverse effects were observed in n=22 (of the entire sample) and n=16 (of the participants without hospital admissions/home treatment).
(b) Educational attainment was dichotomized into two categories. Missing or unknown was classified as less than university attainment.

OR: Odds ratio. CI: Confidence intervals.

**Appendix 4B. Association between psychosocial factors at work and severe adverse effects (a) after COVID-19 vaccines: Multiple logistic regression analysis** **of participants without hospital admissions/home treatment (N=683).**

|  | **Model 1** | | | **Model 2** | | | **Model 3** | | |
| --- | --- | --- | --- | --- | --- | --- | --- | --- | --- |
|  | OR | 95% CI | p | OR | 95% CI | p | OR | 95% CI | p |
| Job demand | 0.94 | 0.76 - 1.17 | 0.590 | 0.95 | 0.73 - 1.24 | 0.708 | 1.02 | 0.77 - 1.35 | 0.884 |
| Job control | 0.93 | 0.73 - 1.18 | 0.544 | 0.94 | 0.71 - 1.24 | 0.651 | 0.94 | 0.7 - 1.26 | 0.671 |
| Coworker support | 0.82 | 0.61 - 1.11 | 0.198 | 0.90 | 0.64 - 1.27 | 0.546 | 0.98 | 0.67 - 1.42 | 0.912 |
| Supervisor support | 1.42 | 1.05 - 1.90 | 0.022* | 1.30 | 0.92 - 1.84 | 0.133 | 1.24 | 0.88 - 1.77 | 0.224 |
| Gender(0: men, 1: women) |  |  |  | 2.54 | 0.65 - 9.84 | 0.179 | 3.91 | 0.89 - 17.27 | 0.072 |
| Age |  |  |  | 0.92 | 0.86 - 0.99 | 0.025* | 0.92 | 0.86 - 0.99 | 0.033* |
| Education (0: less than university, 1: university+) (b) |  |  |  | 0.67 | 0.17 - 2.6 | 0.561 | 0.72 | 0.19 - 2.82 | 0.639 |
| Marital status (0: single, 1: married) |  |  |  | 1.92 | 0.52 - 7.1 | 0.326 | 2.10 | 0.56 - 7.87 | 0.273 |
| Non-manual workers (ref: managers) |  |  |  | 0.54 | 0.05 - 6.09 | 0.618 | 0.66 | 0.06 - 7.43 | 0.738 |
| Manual workers (ref: managers) |  |  |  | 0.61 | 0.04 - 8.77 | 0.715 | 0.76 | 0.05 - 11.45 | 0.845 |
| Healthcare workers (ref: managers) |  |  |  | 2.74 | 0.12 - 61.1 | 0.525 | 2.76 | 0.12 - 61.52 | 0.522 |
| Chronic disease (ref: managers) |  |  |  | 0.94 | 0.11 - 8.21 | 0.958 | 1.15 | 0.12 - 10.65 | 0.903 |
| Vaccination (ref: first time) |  |  |  | 59885447.66 |  | 0.998 | 59191849.53 |  | 0.998 |
| Anxiety about side effects of vaccination |  |  |  |  |  |  | 0.65 | 0.3 - 1.4 | 0.271 |
| Fear and worry about COVID-19 |  |  |  |  |  |  | 0.75 | 0.44 - 1.27 | 0.280 |
| Psychological distress |  |  |  |  |  |  | 1.02 | 0.91 - 1.15 | 0.747 |

(a)	Severe adverse effect was defined as self-reported severe reactions requiring medical care after vaccination (e.g., anaphylaxis). Severe adverse effects were observed in n=22 (of the entire sample) and n=16 (of the participants without hospital admissions/home treatment).

(b)	Educational attainment was dichotomized into two categories. Missing or unknown was classified as less than university attainment.

OR: Odds ratio. CI: Confidential intervals.
